# Does Parental Migration Affect a Child’s Immunization Coverage? A Cross-sectional Analytical Study of India

**DOI:** 10.64898/2026.05.14.26353222

**Authors:** Pritu Dhalaria, Pawan Kumar, Sanjay Kapur, Ajay Kumar Verma, Ajeet Kumar Singh, Pretty Priyadarshini, Kapil Singh, Bhupendra Tripathi, Arindam Ray

**Affiliations:** Immunization Technical Support Unit, Ministry of Health & Family Welfare, Government of India, New Delhi,110070, India; Immunization Division, Ministry of Health & Family Welfare, New Delhi 110011, India; John Snow India, New Delhi, 110070, India; Gates Foundation, New Delhi, 110067, India

**Keywords:** Parental Migration, Immunization Coverage, Determinants, Intervention, IA2030

## Abstract

**Introduction:** India’s immunization initiatives are among the largest globally, characterized by a substantial birth cohort of 27 million children annually, and have achieved significant progress in increasing coverage through the UIP. However, there are still challenges that persist, and multiple determinants contribute to the existing challenges; parental migration is one of them. Migration has always been a key driver of socio-economic and demographic changes, particularly in low and middle-income countries (LMICs). Specifically, there is a need to better understand the vulnerabilities of immunization among recent migrants. To examine this, the study explores the association between a mother’s recent migration and the full immunization coverage of children aged 12-23 months in India.

**Data & Methods:** Our study utilized data from the National Family Health Survey-5 (2019-21). The outcome variable of interest in this study is the receipt of all basic vaccinations (full immunization) for children. The primary predictor variable in this study is the children’s migration status. We used a series of multivariate logistic regression models to examine the relationship between full Immunization and recent migration of children, with some data restrictions in the models.

**Results:** The results show a 17% difference in full immunization between migrant and non-migrant children. The odds ratios for children who had recently migrated were lower for full immunization (OR: 0.39, 95% CI: 0.35–0.43) compared to children who had not recently migrated. Even across the household wealth quintile and social groups, the recent migration of children was associated with being less likely to be fully immunized among children 12-23 months.

**Conclusion:** The findings of this study provide significant quantitative evidence that recent migration (less than 3 years) of children is a key factor influencing Immunization coverage and is a predictor of full vaccination among children aged 12-23 months in India. The recent migration was consistently linked to a lower likelihood of full immunization coverage across different household wealth levels and social groups. This study suggests that recently migrated children are a vulnerable subgroup of the population at risk of not receiving all basic vaccinations by their first birthday.

## Introduction

The Immunization Agenda (IA) 2030 promotes equitable access to full immunization for all, irrespective of location, age, gender, or socioeconomic barriers [1]. It recognizes vaccinations as the most effective public health interventions to safeguard children’s health against disease. Vaccinations advance global health equity by reducing inequalities between high-income and socioeconomically disadvantaged communities [2,3]. Since the introduction of the Expanded Programme on Immunization (EPI) in 1974, vaccinations have prevented 154 million deaths worldwide. This includes 146 million deaths among children under five, with 101 million of these deaths occurring among children less than one year old. These childhood survival gains have occurred across all WHO regions [4]. Estimates suggest that childhood vaccination programs in low- and middle-income countries (LMICs) for ten pathogens from 2000 to 2030 will prevent approximately 69 million deaths. Of these, 37 million deaths were avoided between 2000 and 2019, compared to scenarios without vaccination. Improved worldwide coverage and new vaccine developments could reduce life-time mortality for the 2019 birth cohort by up to 72% [5].

India’s immunization initiatives are among the largest globally, catering to the world’s largest birth cohort of approximately 26 million children and 29 million pregnant women annually [6]. The FIC has improved over recent decades, from 35% in 1992-93 to 76% in 2019-21 [7,8]. With 76.4% full immunization coverage (FIC) and 20% of children partially immunized, India is on track to achieve the global target of the Immunization Agenda 2030 (IA 2030) [9]. Despite India’s efficient immunization system, a significant number of children remain under vaccinated. Coverage drops have been observed across all multidose childhood vaccines, highlighting the persistent challenge of vaccine completion beyond the first dose and substantial gaps and inequalities across regions and socioeconomic groups [10]. Factors like socioeconomic status, maternal education, limited awareness, vaccine hesitancy, and gender bias are well recognized contributors to incomplete vaccination coverage in India. Additional challenges, such as difficult terrain and strained health infrastructure, further compound the issue [7,9,11–13].

Among the various reasons for partial or non-immunization, migration stands out as a significant barrier to immunization uptake. Migrant populations represent some of the most vulnerable groups that face adverse health outcomes and barriers to healthcare services, including childhood immunization [14]. Suboptimal vaccination coverage is a significant concern for migrant populations globally, with implications for both individual and public health. In European countries, evidence suggests that vaccination coverage among both migrant adults and children is markedly lower, around 50%, compared to the native population [15]. Consistently lower coverage in migrant groups is not merely an individual health issue. It is also linked to vaccine-preventable disease (VPD) outbreaks, including measles, varicella, mumps, rubella, and hepatitis A [16]. Similar patterns are observed in the Middle East and North Africa (MENA) region. Data from 15 countries in this region show that routine childhood vaccination coverage can be as low as 36% in certain countries, such as Lebanon, Morocco, and Sudan [17]. Specifically in India, studies from certain regions have also documented low full vaccination coverage among children of migrant construction workers, ranging from 26% to 30% [18,19]. According to the recent Periodic Labour Force Survey (PLFS) 2020-21 estimates, India’s migration rate is approximately 30%. This rate shows a variation between regions, with 26.5% in rural areas and 34.9% in urban areas [20]. This significant demographic shift, in turn, leads many people to establish new living arrangements in urban slums and peri-urban areas. This transition has the potential to affect their access to and utilization of health services, including routine immunization of infants and children. Achieving full immunization coverage is a continuous, year-long process that requires multiple visits, as outlined in the National Immunization Schedule. Therefore, migration during this critical period can lead to dropouts and delays in immunization uptake.

Migration serves as a significant catalyst for socio-economic and demographic shifts, particularly in LMICs. The association between migration and health outcomes is diverse and is influenced by several factors. These include the characteristics of the migrant, the stage of life at which the migration occurs, the specific origins and destinations involved, and the underlying motivations for migrating [14,21,22].

### Conceptual framework

Access to healthcare services is a major concern in social science and public health research for vulnerable populations. The utilization of health services, including immunization, by migrants is commonly analyzed through three theoretical perspectives: migrant disruption, adaptation, and assimilation [14,23,24]. The disruption perspective holds that migration disrupts the natural progression of demographic events in migrants’ lives, thereby impacting health behaviors and outcomes. Disruption is assumed to bring short-term behavioral changes. This is often due to insecurity in livelihoods, unmet basic needs, and vulnerabilities in existing health beliefs, practices, and social norms. Conversely, the adaptation hypothesis proposes that migrants intentionally, although temporarily, modify their behavior to conform to the prevailing social norms of their new environment. The assimilation hypothesis suggests that migrants progressively internalize the norms of the host society before fully integrating into the community [25–28].

The literature identifies several dimensions related to the root causes of vaccination coverage gaps and vaccine uptake, often categorized into key themes. The evidence-informed 4As-Access, Affordability, Awareness, and Acceptance Taxonomy is a prominent theoretical framework for determining vaccine uptake behavior in childhood and adults [29]. This demonstrates that factors such as livelihood insecurity, health beliefs, behavioral vulnerability, and social norms collectively influence vaccination access and outcomes. This complex interplay directly influences the consistent uptake of routine vaccines and their coverage. Language, literacy, and communication barriers emerged as critical factors that can reduce vaccine uptake [30]. Studies indicate that migrants often face challenges accessing public health services, including child vaccination. Beyond financial access concerns, non-financial barriers also impede vaccination, even among caregivers who intend to immunize their children. These included opportunity costs, competing priorities, and rigid session timings and schedules [29,31]. Information and perception-related issues contribute to hesitancy, often stemming from a lack of or exposure to misinformation, a low perception of the disease’s risk or the importance of vaccination, and the absence of an explicit physician or frontline health worker recommendation. Thus, the 4As taxonomy helps classify and describe different potential causes of lower vaccination uptake amongst migrants. Building on this framework, our study hypothesizes that recent migrants may be less likely than settled migrants and the native population to complete their full immunization (see Figure 1). The hypothesis has been translated into a flow diagram to better portray the effects of migration on immunization uptake.

**Figure 1.**
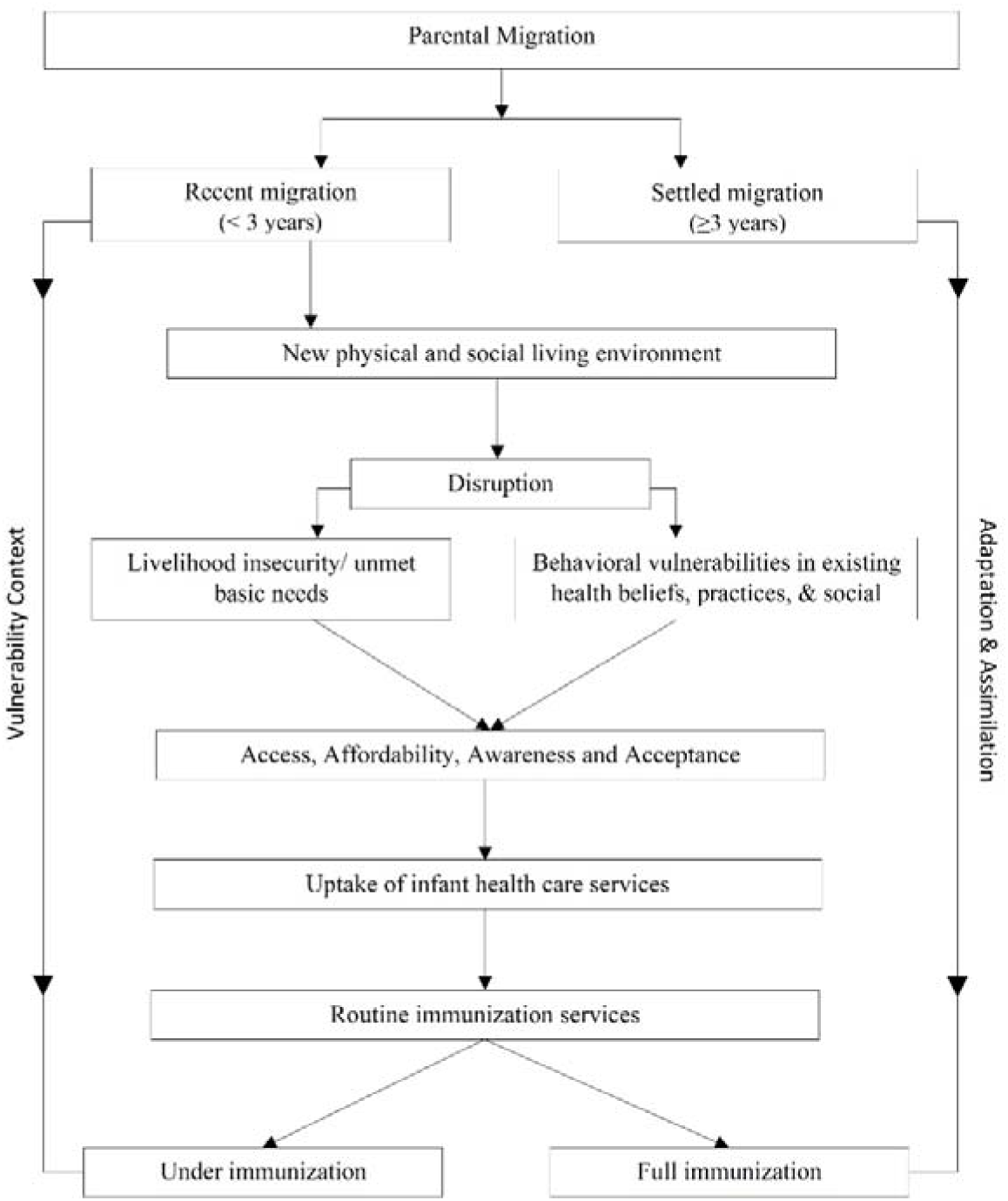
Hypothesized theoretical framework of how migration influences immunization uptake among children.

**Figure 2.**
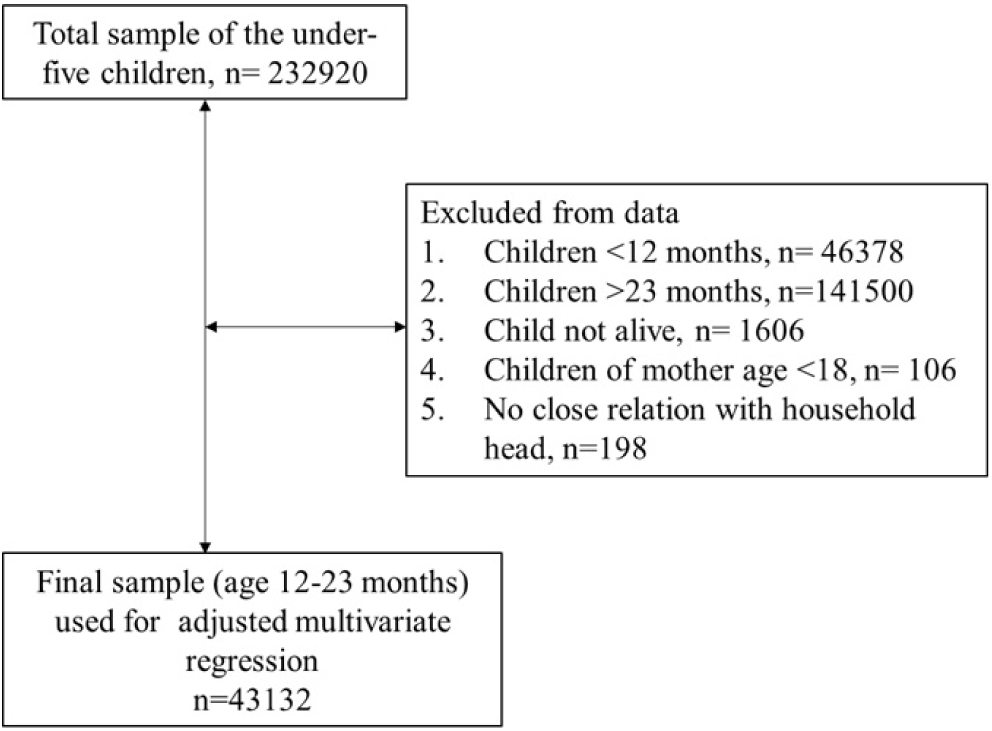
Flow chart of the analytical sample, NFHS 2019–21.

Understanding the immunization status of migrant children is challenging due to the complex nature of migration. There are very few studies in India that have examined the relationship between migration and children’s immunization status. Existing nationally representative studies show inconsistent findings regarding the association between migration status and full immunization coverage. Notably, no studies have specifically analyzed the relationship between short-term migration and full immunization for children aged 12-23 months using a nationally representative sample. Two studies on migration and childhood immunization exist, but both have statistical and conceptual limitations. Prusty and Keshri (2015) investigated the association between child nutrition and immunization among migrants and non-migrants using urban sample data from the National Family Health Survey (NFHS) 2005-06 [32]. Mishra et al. (2020) examined migration across socioeconomic groups and immunization coverage using NFHS 2015-16 data [33]. Their findings suggested that migrated children (< 4 years) had better vaccination coverage and more likely to vaccine uptake compared to non-migrants. Some state and city-specific studies indicate that immunization coverage among migrant children was either found to be low or no significant difference compared to non-migrants and settled migrants, depending on the location [19,28,34]. Conceptually, these studies did not clearly distinguish recent migrants from other groups, often treating all periods of residence as migration.

Recent migrants are often primarily focused on meeting their daily livelihood needs and tend to remain socially isolated compared to settled migrants and native populations. Unlike long-term residents, new arrivals often lack established local support networks to help them navigate the healthcare system or access information about available services. This challenge is further compounded by poor living and housing conditions and limited access to essential healthcare. In line with the goals of Immunization Agenda 2030 (IA2030) and its commitment to “leave no child behind,” comprehensive tracking of children is essential; however, migrant children remain one of the most difficult groups to reach across the stratum.

Migration is a major and growing phenomenon in developing countries such as India, yet its implications for child health, particularly immunization, remain insufficiently understood. Our study aims to examine the relationship between recent parental migration and children’s complete immunization status aged 12–23 months using the most recent nationally representative data. There is a clear gap in the literature regarding the nuanced stratification of migrant populations, especially in distinguishing between recent and settled migrants and non-migrant groups, and in assessing differences in immunization coverage across these categories. The duration of stay is a critical but often overlooked dimension of migration dynamics, as recent migrants are likely to be the most vulnerable and have lower uptake of public health services, including child immunization, compared to other groups. By addressing this gap, the study seeks to generate deeper insights into the specific vulnerabilities migrant populations face in accessing childhood immunization services. These findings can help identify high-risk districts and clusters with significant migrant populations, enabling more targeted, context-sensitive interventions. Such evidence is essential for strengthening programmatic strategies and accelerating progress toward equitable immunization coverage in line with IA2030 goals.

## Methodology

### Data Sources

The study is based on data from the National Family Health Survey (NFHS-5) 2019-21. NFHS-5 is a nationally representative survey conducted under the Ministry of Health and Family Welfare (MoHFW), Government of India. The primary objective of NFHS 2019-21 was to collect high-quality data on health and family welfare indicators, enabling policymakers to track progress in India’s health sector and identify gaps in specific health programs. The NFHS data provides information on maternal and child health, including fertility, infant and child mortality, maternal health, and uptake of healthcare services. The sample size of NFHS-5 was designed to generate indicators at district, state, and union territory (UT) levels, covering 707 districts, 28 states, and 8 UTs. NFHS-5 employed a stratified two-stage sampling method to ensure representativeness, operational feasibility, and cost-efficiency. In the first stage, the Indian Census provided the sampling frame for selecting primary sampling units (PSUs) in rural areas and census enumeration blocks (CEBs) in urban areas. In the second stage, 22 PSUs and CEBs were randomly selected in each district. 724,115 women aged 15-49 from 636,699 chosen households were interviewed. We included 43,132 children aged 12-23 months to meet our study goals.

### Outcome variable

The outcome variable of interest in this study is the coverage of all basic vaccinations (fully immunized) for children. A child is considered fully immunized when they have received a dose of BCG, three doses of DPT and Polio each, and one dose of measles vaccine at any time before the survey, as documented on the vaccination card or by direct reporting by the mother.

### Predictor variable

The primary predictor variable in this study is children’s migration status. NFHS-5 does not provide direct information on the respondents’ migration status to their previous place of residence. Indeed, NFHS-5 inquiries about how long the respondents, the children’s mothers, have been residing at their current address, offering three answer choices: ‘years in numbers’, ‘always’, and ‘visitors’. We classified recent migration as living at the current residence for less than 3 years, including visitors with a relationship to the household, such as a daughter, daughter-in-law, or grandchild. It is assumed that these movements result in minimal alteration to the physical or social environment concerning healthcare access and behavior. Close relatives (visitors) of the household are also included in the scope of temporary childbirth migration. This is because they benefit from additional care and support, are expected to have reduced workload at the natal home, and experience lower health-related costs.

We included the child and mother’s socio-demographic background variables such as gender of child, mother’s age at childbirth (15-19 years, 20-29 years and 30+ years), maternal education in years (no schooling, <5 years complete, 5-8 years complete, 9-12 years, and more >12 years), media exposure at least once in a week (No, Yes), child birth order (1^st^ order, 2^nd^, 3^rd^, 4^th^ & more orders), institutional delivery (Public health facility, Private health facility, No health facility), Intended birth (Yes, No), current place of residence (Urban and Rural), social groups (Schedule caste, Schedule tribe, Other backward caste and Others), wealth quintile (Poorest, Poorer, Middle, Richer, and Richest), religion (Hindu, Muslim, Christian, Sikh and Others), regions of India (North, Central, East, Northeast, West and South).

### Statistical analysis

Initially, our study analyzed the distribution of the sample in relation to the outcome and predictor variables, along with various socio-demographic factors. Next, we estimated the inequality in full immunization by recent migration status across 36 Indian states/UTs, along with several socio-economic variables, including wealth quintile, maternal education, and social groups. We used a series of multivariate logistic regression models to examine the relationship between full immunization and recent migration (Yes/No), while accounting for data restrictions. Furthermore, we created two additional supplementary models of recent migration, one for migration of less than 3 years and permanent residence, and another for migration of less than 3 years and settled migration (migration of 3 years or more), to examine the relationship between full immunization and recent migration as a sensitivity analysis. Finally, a series of multivariate logistic regression models was employed to examine the relationship between full immunization and recent migration across different wealth quintiles and social groups, aiming to demonstrate the interaction effect and address heterogeneity.

## Results

The distribution of the study sample is shown in Table 1. The table indicates that 76.6% of participants reported being fully immunized, while 23.4% reported being under-immunized, including children who had received at least one vaccination. Additionally, 9.6% of the sample reported recent migration within the past three years, including those who were visitors at their current residence, while 90.4% reported no migration, having settled in their current residence and maintained permanent residence at the time of the survey.

**Table 1:**
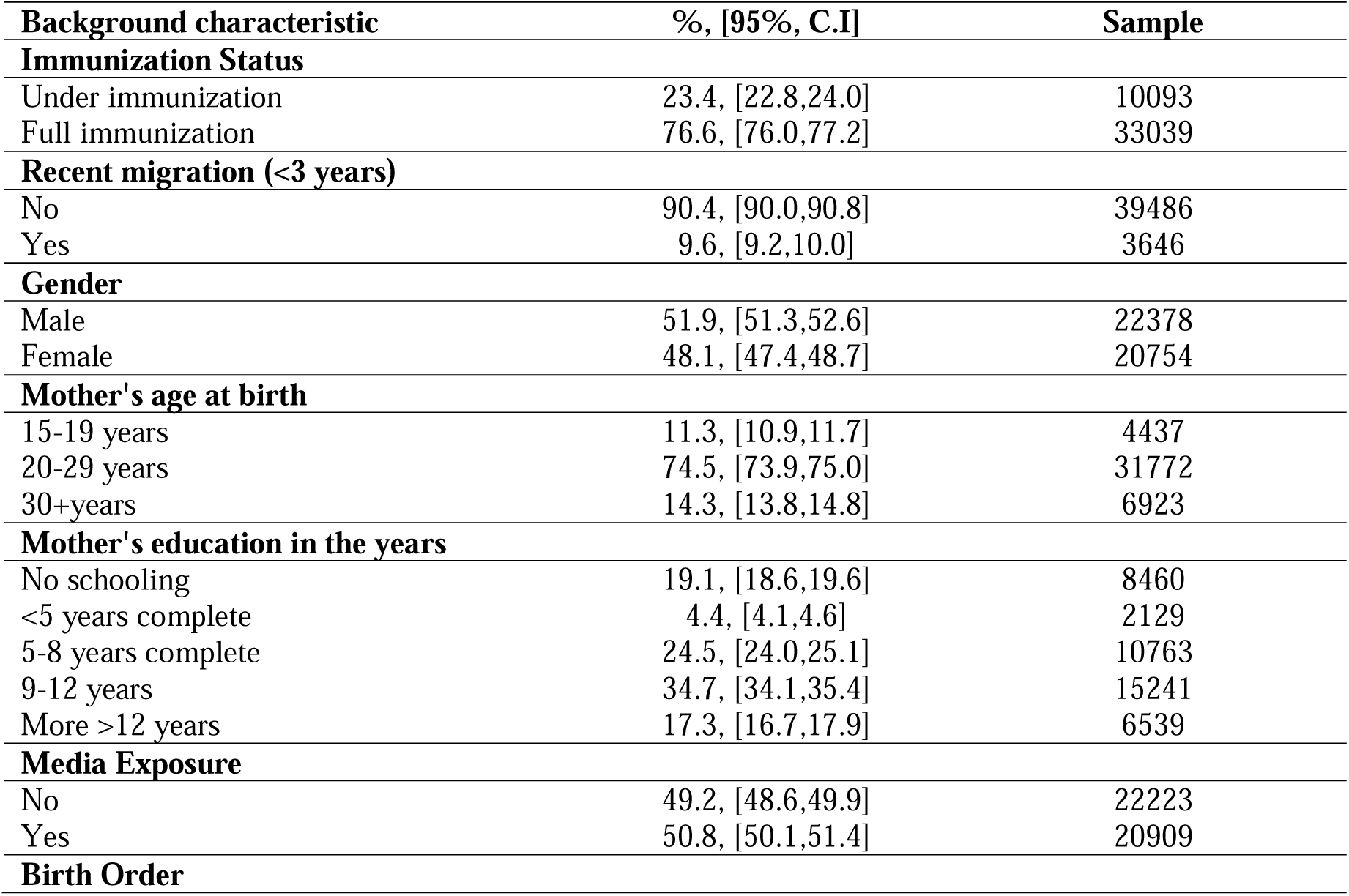
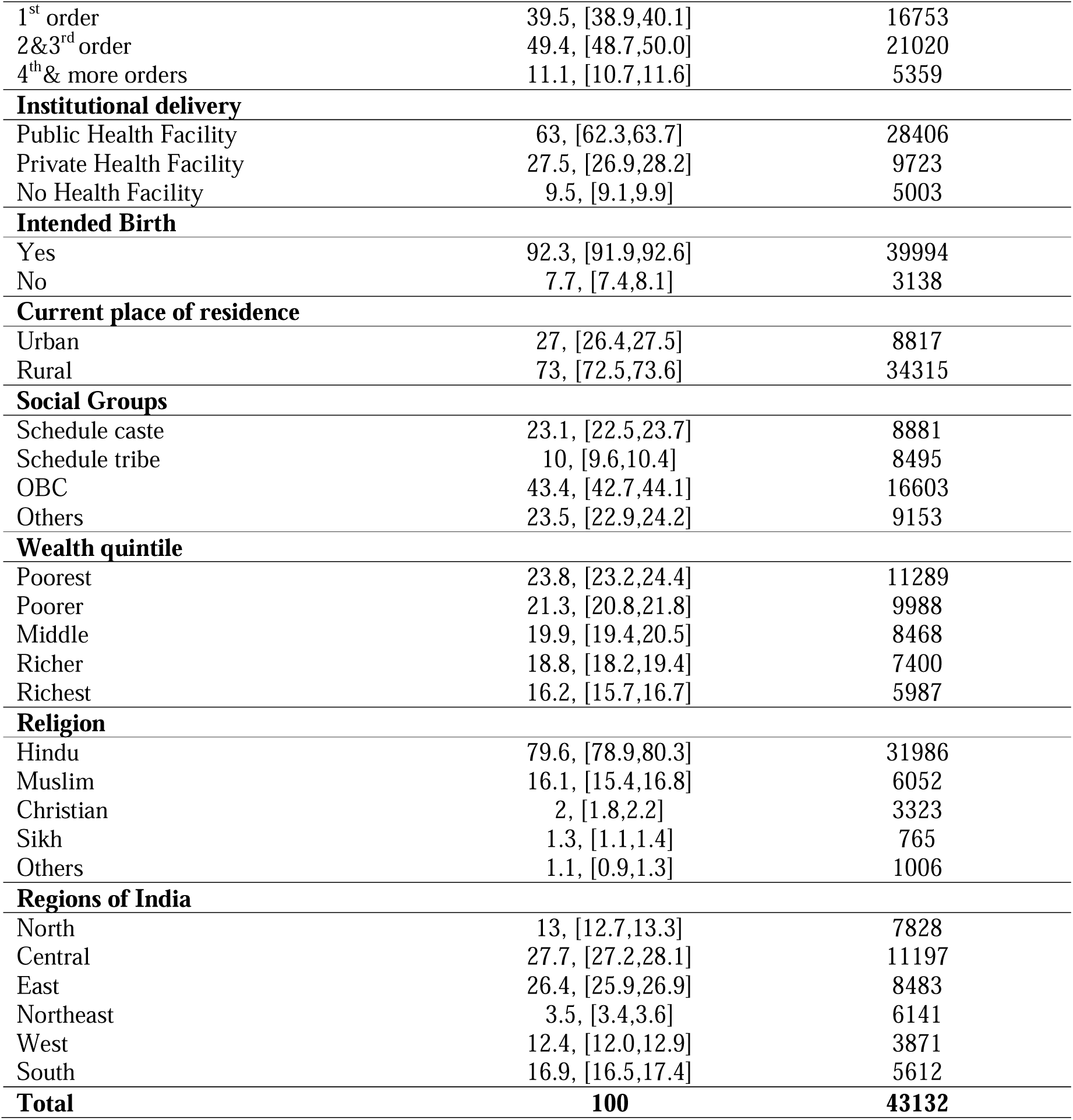
Characteristics of the indicators, India, 2019-21.

| **Background characteristic** | **%, [95%, C.I]** | **Sample** |
| --- | --- | --- |
| **Immunization Status** |  |  |
| Under immunization | 23.4, [22.8,24.0] | 10093 |
| Full immunization | 76.6, [76.0,77.2] | 33039 |
| **Recent migration (<3 years)** |  |  |
| No | 90.4, [90.0,90.8] | 39486 |
| Yes | 9.6, [9.2,10.0] | 3646 |
| **Gender** |  |  |
| Male | 51.9, [51.3,52.6] | 22378 |
| Female | 48.1, [47.4,48.7] | 20754 |
| **Mother's age at birth** |  |  |
| 15-19 years | 11.3, [10.9,11.7] | 4437 |
| 20-29 years | 74.5, [73.9,75.0] | 31772 |
| 30+years | 14.3, [13.8,14.8] | 6923 |
| **Mother's education in the years** |  |  |
| No schooling | 19.1, [18.6,19.6] | 8460 |
| <5 years complete | 4.4, [4.1,4.6] | 2129 |
| 5-8 years complete | 24.5, [24.0,25.1] | 10763 |
| 9-12 years | 34.7, [34.1,35.4] | 15241 |
| More >12 years | 17.3, [16.7,17.9] | 6539 |
| **Media Exposure** |  |  |
| No | 49.2, [48.6,49.9] | 22223 |
| Yes | 50.8, [50.1,51.4] | 20909 |
| **Birth Order** |  |  |
| 1<sup>st</sup> order | 39.5, [38.9,40.1] | 16753 |
| 2&3<sup>rd</sup> order | 49.4, [48.7,50.0] | 21020 |
| 4<sup>th</sup>& more orders | 11.1, [10.7,11.6] | 5359 |
| **Institutional delivery** |  |  |
| Public Health Facility | 63, [62.3,63.7] | 28406 |
| Private Health Facility | 27.5, [26.9,28.2] | 9723 |
| No Health Facility | 9.5, [9.1,9.9] | 5003 |
| **Intended Birth** |  |  |
| Yes | 92.3, [91.9,92.6] | 39994 |
| No | 7.7, [7.4,8.1] | 3138 |
| **Current place of residence** |  |  |
| Urban | 27, [26.4,27.5] | 8817 |
| Rural | 73, [72.5,73.6] | 34315 |
| **Social Groups** |  |  |
| Schedule caste | 23.1, [22.5,23.7] | 8881 |
| Schedule tribe | 10, [9.6,10.4] | 8495 |
| OBC | 43.4, [42.7,44.1] | 16603 |
| Others | 23.5, [22.9,24.2] | 9153 |
| **Wealth quintile** |  |  |
| Poorest | 23.8, [23.2,24.4] | 11289 |
| Poorer | 21.3, [20.8,21.8] | 9988 |
| Middle | 19.9, [19.4,20.5] | 8468 |
| Richer | 18.8, [18.2,19.4] | 7400 |
| Richest | 16.2, [15.7,16.7] | 5987 |
| **Religion** |  |  |
| Hindu | 79.6, [78.9,80.3] | 31986 |
| Muslim | 16.1, [15.4,16.8] | 6052 |
| Christian | 2, [1.8,2.2] | 3323 |
| Sikh | 1.3, [1.1,1.4] | 765 |
| Others | 1.1, [0.9,1.3] | 1006 |
| **Regions of India** |  |  |
| North | 13, [12.7,13.3] | 7828 |
| Central | 27.7, [27.2,28.1] | 11197 |
| East | 26.4, [25.9,26.9] | 8483 |
| Northeast | 3.5, [3.4,3.6] | 6141 |
| West | 12.4, [12.0,12.9] | 3871 |
| South | 16.9, [16.5,17.4] | 5612 |
| **Total** | **100** | **43132** |

The study shows disparities in full immunization coverage across different states/UTs in India and among various socio-economic groups, particularly in relation to recent migration. A notable 17% disparity in full immunization rates among children is observed when comparing those with recent migration status, as shown in Figure 3. Among major states, only Odisha (3%) and Assam (1%) showed negligible differences in full immunization coverage between recent migrants and non-migrants. Conversely, Ladakh, Chandigarh, Meghalaya, and Lakshadweep showed lower vaccination rates among children of non-recent migrants than among those of recent migrants. Figure 4 displays full immunization coverage among children from different socio-economic backgrounds, categorized by their recent migration status. The data indicate that children from the poorest households, those whose mothers have less than five years of education, and who are from the Scheduled Castes, have lower immunization rates, especially among those who are recent migrants.

**Figure 3:**
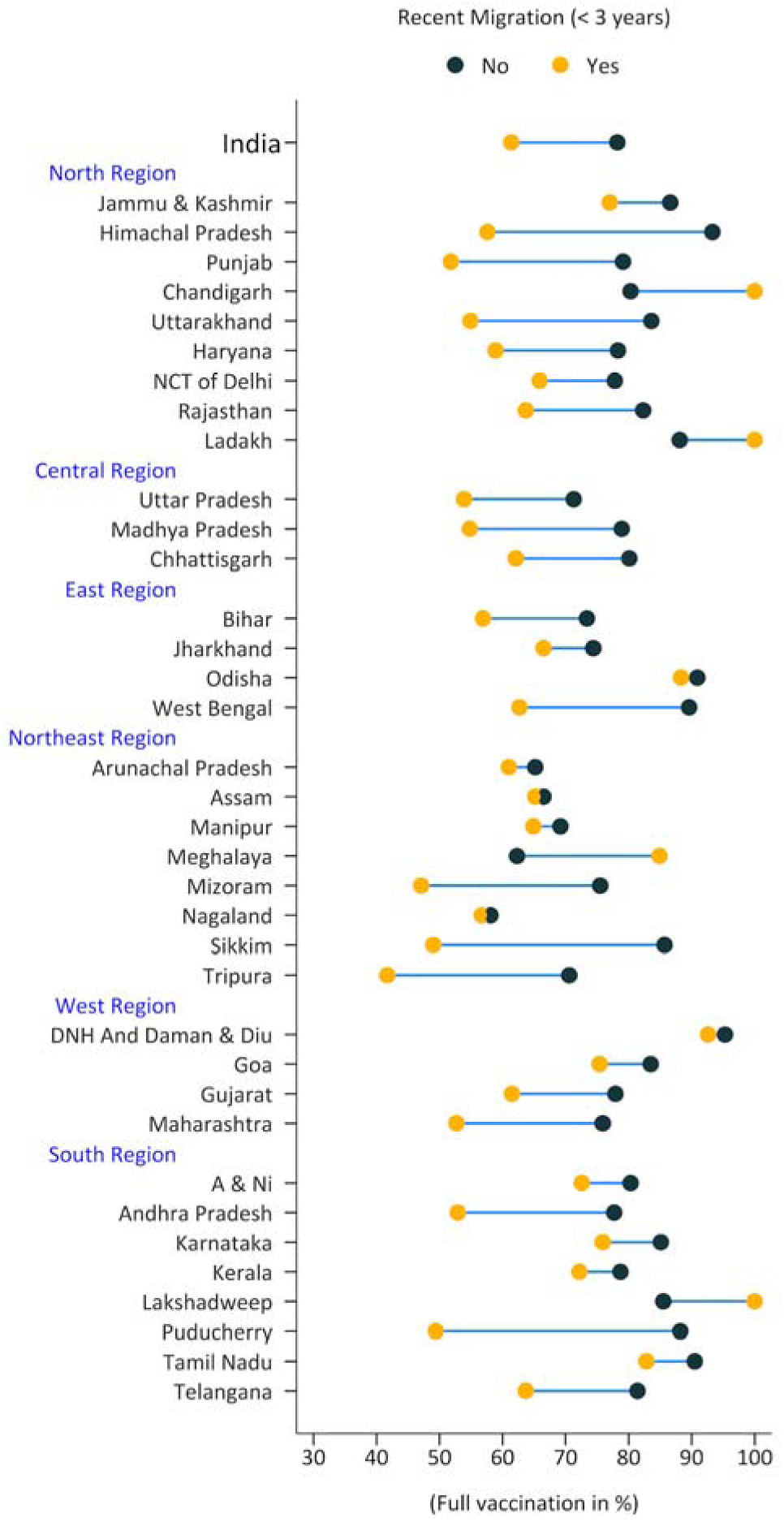
Inequality in all basic vaccinations among children aged 12-23 months by mothers’ recent migration status across States/UTs, India, 2019-21

**Figure 4:**
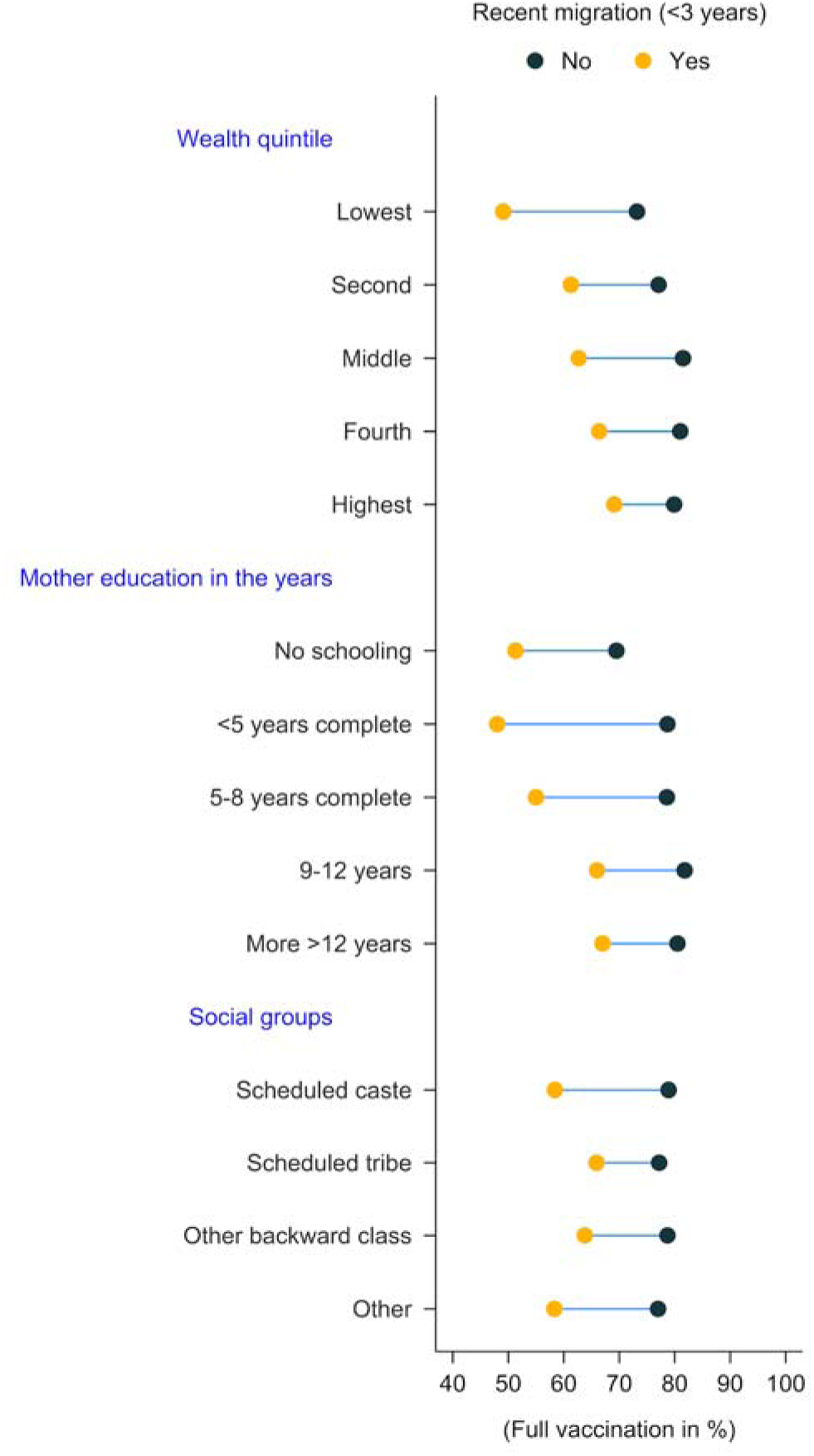
Inequality in all basic vaccinations among children aged 12-23 months by mothers’ recent migration status across different socio-economic backgrounds, mothers’ education status, and social groups in India, 2019-21

Table 2 shows the results of a multivariate logistic regression analysis on full immunization among children aged 12-23 months. In Model I, the odds of full immunization were lower among children who had recently migrated (OR: 0.39, 95% CI: 0.35–0.43) than among those who had not recently migrated. In Model II (limited to children whose mother did not live with their husband and who had multiple wives), the odds ratio for full immunization was lower among recently migrated children (OR: 0.38, 95% CI: 0.34–0.44) than non-migrated children. Furthermore, in Model III (limited to children who were visitors and whose mothers were not living with their husbands and had multiple wives), there was a lower odds ratio for full immunization among recent migrants (OR: 0.52, 95% CI: 0.45–0.62) compared to non-migrants. This pattern suggests a consistently lower likelihood of full immunization among recently migrated children across different analytical models.

**Table 2.** Multivariate logistic regression on Full immunization among children 12-23 months, India, 2019-21

|  | **Model-I** | **Model-II<sup>#</sup>** | **Model-II<sup>##</sup>** |
| --- | --- | --- | --- |
| **Background characteristic** | **OR, [95%, C.I]** | **OR, [95%, C.I]** | **OR, [95%, C.I]** |
| **Recent Migration (<3 years)** |  |  |  |
| No <sup>Ref</sup> | 1 | 1 | 1 |
| Yes | 0.39*** [0.35,0.43] | 0.38*** [0.34,0.44] | 0.52*** [0.45,0.62] |
| **Gender** |  |  |  |
| Male <sup>Ref</sup> | 1 | 1 | 1 |
| Female | 0.96 [0.90,1.03] | 0.96 [0.89,1.03] | 0.97 [0.90,1.04] |
| **Mother's age at birth** |  |  |  |
| 15-19 years <sup>Ref</sup> | 1 | 1 | 1 |
| 20-29 years | 1.17** [1.05,1.31] | 1.15* [1.01,1.30] | 1.14* [1.00,1.30] |
| 30+years | 1.29** [1.10,1.50] | 1.24* [1.05,1.47] | 1.24* [1.04,1.47] |
| **Mother's education in the years** |  |  |  |
| No schooling <sup>Ref</sup> | 1 | 1 | 1 |
| <5 years complete | 1.45*** [1.24,1.69] | 1.44*** [1.22,1.71] | 1.48*** [1.25,1.76] |
| 5-8 years complete | 1.40*** [1.27,1.53] | 1.41*** [1.27,1.56] | 1.42*** [1.28,1.58] |
| 9-12 years | 1.50*** [1.36,1.66] | 1.51*** [1.36,1.69] | 1.52*** [1.36,1.70] |
| More >12 years | 1.30*** [1.13,1.49] | 1.26** [1.08,1.47] | 1.26** [1.07,1.48] |
| **Media Exposure** |  |  |  |
| No <sup>Ref</sup> | 1 | 1 | 1 |
| Yes | 1.18*** [1.09,1.27] | 1.17*** [1.08,1.28] | 1.19*** [1.09,1.30] |
| **Birth Order** |  |  |  |
| 1<sup>st</sup> order <sup>Ref</sup> | 1 | 1 | 1 |
| 2 & 3<sup>rd</sup> order | 0.81*** [0.75,0.87] | 0.80*** [0.73,0.88] | 0.80*** [0.73,0.87] |
| 4<sup>th</sup> & more order | 0.69*** [0.61,0.77] | 0.68*** [0.59,0.78] | 0.67*** [0.58,0.76] |
| **Intended Birth** |  |  |  |
| Yes <sup>Ref</sup> | 1 | 1 | 1 |
| No | 0.91 [0.80,1.04] | 0.88 [0.76,1.02] | 0.89 [0.77,1.02] |
| **Institutional delivery** |  |  |  |
| Public Health Facility <sup>Ref</sup> | 1 | 1 | 1 |
| Private Health Facility | 0.80*** [0.73,0.87] | 0.79*** [0.72,0.86] | 0.79*** [0.72,0.87] |
| No Health Facility | 0.63*** [0.57,0.70] | 0.62*** [0.55,0.69] | 0.61*** [0.55,0.69] |
| **Current place of residence** |  |  |  |
| Urban <sup>Ref</sup> | 1 | 1 | 1 |
| Rural | 1.25*** [1.13,1.38] | 1.28*** [1.15,1.42] | 1.30*** [1.17,1.45] |
| **Social Groups** |  |  |  |
| Schedule caste <sup>Ref</sup> | 1 | 1 | 1 |
| Schedule tribe | 1.09 [0.94,1.25] | 1.07 [0.92,1.25] | 1.06 [0.90,1.24] |
| OBC | 1.03 [0.94,1.12] | 1.04 [0.94,1.14] | 1.05 [0.95,1.16] |
| Others | 0.94 [0.85,1.05] | 0.94 [0.83,1.05] | 0.94 [0.83,1.06] |
| **Wealth quintile** |  |  |  |
| Poorest <sup>Ref</sup> | 1 | 1 | 1 |
| Poorer | 1.14** [1.04,1.24] | 1.10 [1.00,1.22] | 1.10 [0.99,1.21] |
| Middle | 1.34*** [1.21,1.49] | 1.32*** [1.18,1.49] | 1.30*** [1.15,1.46] |
| Richer | 1.35*** [1.19,1.55] | 1.35*** [1.16,1.56] | 1.29*** [1.11,1.50] |
| Richest | 1.38*** [1.18,1.62] | 1.43*** [1.21,1.68] | 1.38*** [1.17,1.63] |
| **Religion** |  |  |  |
| Hindu <sup>Ref</sup> | 1 | 1 | 1 |
| Muslim | 0.80*** [0.73,0.89] | 0.80*** [0.71,0.89] | 0.79*** [0.71,0.89] |
| Christian | 1.17 [0.95,1.45] | 1.12 [0.89,1.41] | 1.16 [0.93,1.45] |
| Sikh | 0.85 [0.67,1.08] | 0.84 [0.66,1.09] | 0.83 [0.65,1.08] |
| Others | 1.33 [0.88,2.02] | 1.42 [0.94,2.15] | 1.37 [0.91,2.06] |
| **Regions of India** |  |  |  |
| North <sup>Ref</sup> | 1 | 1 | 1 |
| Central | 0.73*** [0.66,0.80] | 0.74*** [0.66,0.82] | 0.72*** [0.65,0.81] |
| East | 1.17** [1.04,1.31] | 1.20** [1.06,1.36] | 1.16* [1.02,1.32] |
| Northeast | 0.53*** [0.46,0.60] | 0.52*** [0.45,0.59] | 0.50*** [0.43,0.57] |
| West | 0.75*** [0.64,0.87] | 0.77** [0.66,0.90] | 0.76*** [0.65,0.89] |
| South | 1.12 [0.98,1.27] | 1.10 [0.96,1.26] | 1.07 [0.92,1.23] |
| **Sample** | **43132** | **37172** | **36357** |
| Note-Ref=Reference; p value * p<0.05, ** p<0.01, *** p<0.001; # Restricted to women who were not living with their husbands and whose husbands had more than one wife. ## Restricted to visitors + # |  |  |  |

Table 3 shows the results of a multivariate logistic regression analysis on full immunization in relation to children’s recent migration and permanent residence at their current residential place. The analysis found that children who had been recently migrated (less than 3 years ago) had lower odds of being fully immunized than children of permanent residents (OR: 0.67, 95% CI: 0.54–0.83). In Model II of Table 3 (limited to children of mothers not living with their husbands or husbands with additional wives), the results still indicated lower odds for full immunization (OR: 0.70, 95% CI: 0.53–0.92) among children with recent migration compared to those who always resided at their current location. We further limited the analysis to children whose mothers were not living with their husbands or whose husbands had additional wives (Model II of Table 3). The findings were consistent, as children with recent migration had lower odds of full immunization (OR: 0.70, 95% CI: 0.53–0.92) compared to those who had always resided at their current location.

**Table 3.** Multivariate logistic regression on full immunization among children 12-23 months by mother’s recent migration and permanent residences, India, 2019-21

|  | **Model-I** | **Model-II<sup>#</sup>** |
| --- | --- | --- |
| **Background characteristic** | **OR, [95%, C.I]** | **OR, [95%, C.I]** |
| **Recent Migration (<3 years)** |  |  |
| No <sup>Ref</sup> | 1 | 1 |
| Yes | 0.67*** [0.54,0.83] | 0.70* [0.53,0.92] |
| **Gender** |  |  |
| Male <sup>Ref</sup> | 1 | 1 |
| Female | 1.14 [0.94,1.38] | 1.22 [0.96,1.54] |
| **Mother's age at birth** |  |  |
| 15-19 years <sup>Ref</sup> | 1 | 1 |
| 20-29 years | 1.42* [1.07,1.88] | 1.29 [0.90,1.86] |
| 30+years | 1.45 [0.95,2.22] | 1.26 [0.75,2.11] |
| **Mother's education in the years** |  |  |
| No schooling <sup>Ref</sup> | 1 | 1 |
| <5 years complete | 1.34 [0.85,2.12] | 1.48 [0.85,2.58] |
| 5-8 years complete | 1.24 [0.91,1.68] | 1.40 [0.96,2.02] |
| 9-12 years | 1.71** [1.24,2.36] | 1.69** [1.14,2.51] |
| More >12 years | 1.30 [0.89,1.89] | 1.27 [0.80,2.02] |
| **Media Exposure** |  |  |
| No <sup>Ref</sup> | 1 | 1 |
| Yes | 1.03 [0.81,1.31] | 1.07 [0.81,1.43] |
| **Birth Order** |  |  |
| 1<sup>st</sup> order <sup>Ref</sup> | 1 | 1 |
| 2 & 3<sup>rd</sup> order | 0.77* [0.61,0.96] | 0.78 [0.59,1.03] |
| 4<sup>th</sup> & more order | 0.56** [0.37,0.86] | 0.57* [0.34,0.94] |
| **Intended Birth** |  |  |
| Yes <sup>Ref</sup> | 1 | 1 |
| No | 1.03 [0.74,1.44] | 1.06 [0.71,1.57] |
| **Institutional delivery** |  |  |
| Public Health Facility <sup>Ref</sup> | 1 | 1 |
| Private Health Facility | 0.88 [0.69,1.12] | 0.95 [0.71,1.28] |
| No Health Facility | 0.59** [0.42,0.83] | 0.49*** [0.33,0.71] |
| **Current place of residence** |  |  |
| Urban <sup>Ref</sup> | 1 | 1 |
| Rural | 0.91 [0.72,1.15] | 0.94 [0.72,1.23] |
| **Social Groups** |  |  |
| Schedule caste <sup>Ref</sup> | 1 | 1 |
| Schedule tribe | 1.29 [0.89,1.86] | 1.38 [0.89,2.13] |
| OBC | 1.09 [0.84,1.40] | 1.15 [0.84,1.59] |
| Others | 0.87 [0.62,1.21] | 0.80 [0.53,1.21] |
| **Wealth quintile** |  |  |
| Poorest <sup>Ref</sup> | 1 | 1 |
| Poorer | 1.41* [1.06,1.87] | 1.30 [0.91,1.87] |
| Middle | 1.26 [0.91,1.75] | 1.18 [0.79,1.75] |
| Richer | 1.18 [0.82,1.70] | 1.09 [0.70,1.69] |
| Richest | 1.34 [0.86,2.09] | 1.29 [0.76,2.17] |
| **Religion** |  |  |
| Hindu <sup>Ref</sup> | 1 | 1 |
| Muslim | 1.18 [0.87,1.60] | 1.37 [0.93,2.02] |
| Christian | 1.47 [0.91,2.38] | 1.62 [0.94,2.80] |
| Sikh | 1.01 [0.49,2.08] | 1.06 [0.44,2.57] |
| Others | 1.94 [0.77,4.89] | 6.00*** [2.68,13.46] |
| **Regions of India** |  |  |
| North <sup>Ref</sup> | 1 | 1 |
| Central | 0.69* [0.50,0.96] | 0.74 [0.50,1.10] |
| East | 0.97 [0.69,1.37] | 0.96 [0.63,1.47] |
| Northeast | 0.48** [0.30,0.79] | 0.40*** [0.23,0.68] |
| West | 0.73 [0.48,1.10] | 0.76 [0.48,1.21] |
| South | 1.32 [0.92,1.90] | 1.28 [0.85,1.94] |
| **Sample** | **4763** | **3581** |
| Note- * p<0.05, ** p<0.01, *** p<0.001; # Restricted to women who were not living with their husbands and whose husbands had more than one wife. |  |  |

Table 4 presents a multivariate logistic regression analysis of full immunization in relation to recent migration and settled migration (living at the current location for 3 or more years but not permanently residing there) among children aged 12-23 months. In Model I, the odds ratio for full immunization was lower among recently migrated children (OR: 0.52, 95% CI: 0.46–0.60) compared with settled migrant children. In Model II (restricted to children whose mothers do not live with their husbands and have husbands with additional wives), the odds ratio for full immunization was lower (OR: 0.56, 95% CI: 0.48–0.66) among recently migrated children than among children with settled migration.

**Table 4.**
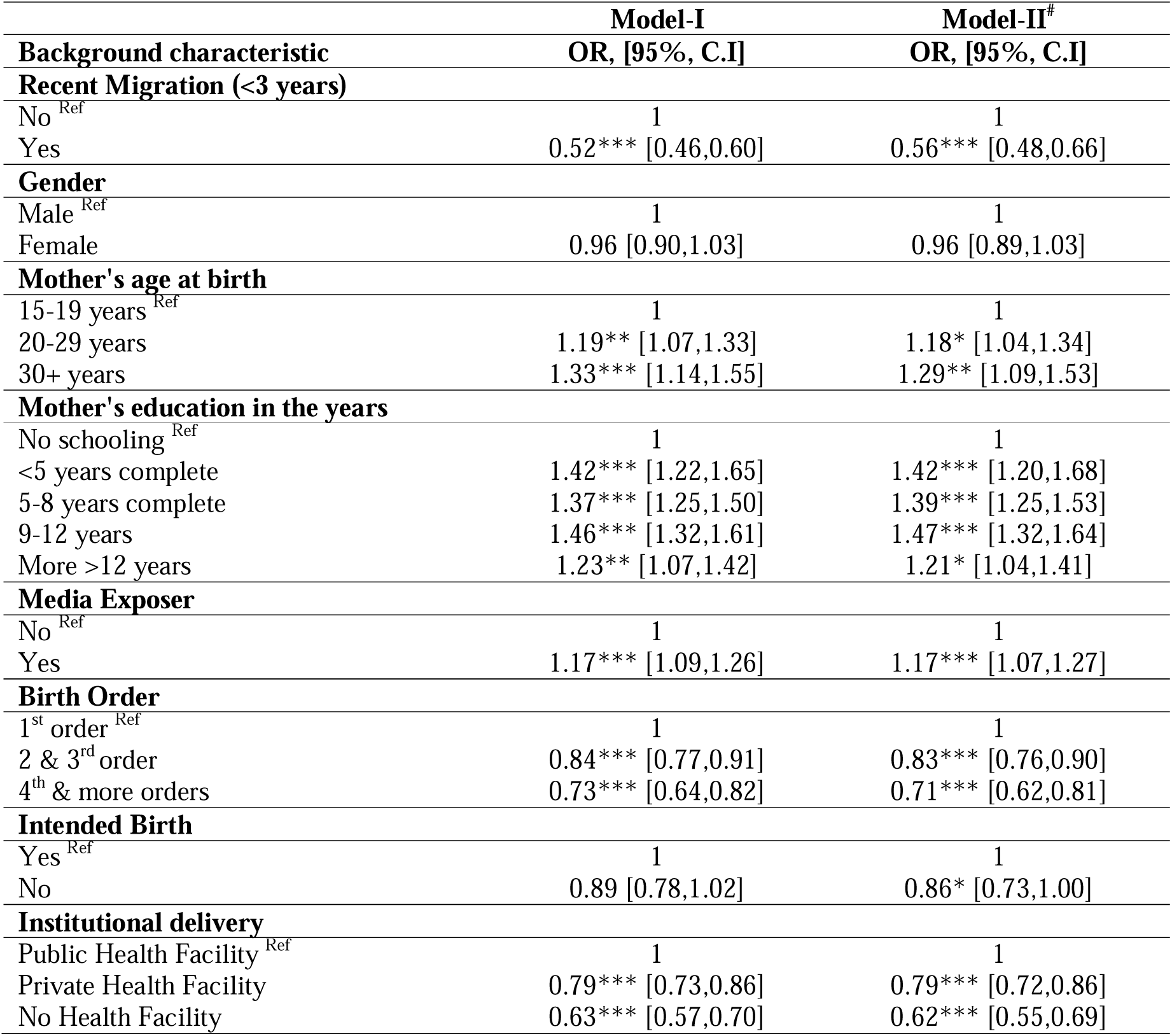
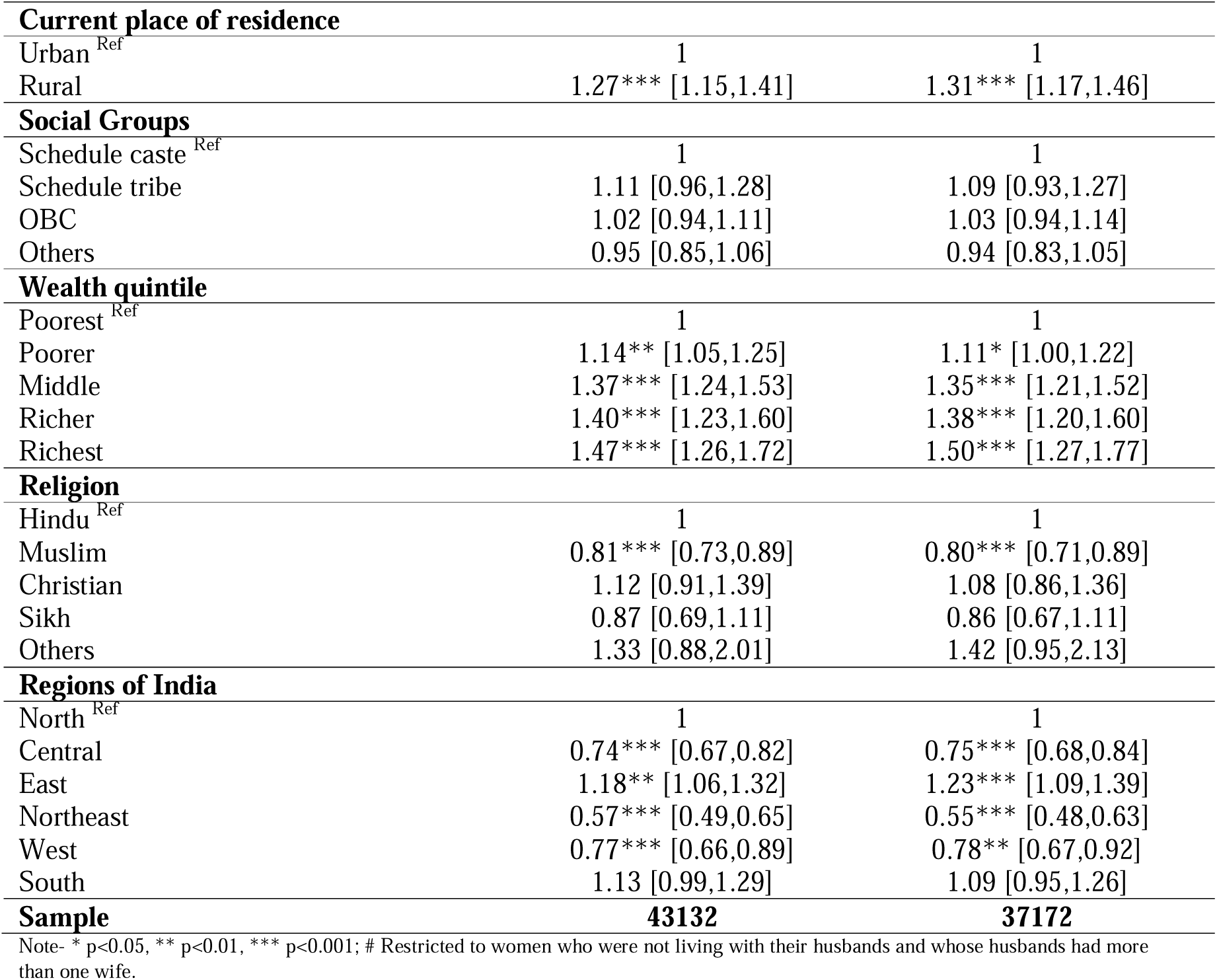
Multivariate logistic regression on Full immunization among children 12-23 months by mother’s recent migration and settled migration, India, 2019-21

Table 5 presents the results of a multivariate logistic regression examining full immunization among children with migrant status, stratified by household wealth. The analysis reveals that recently migrated children across all household wealth quintiles had significantly lower odds of being fully immunized than their non-migrant counterparts. Specifically, the odds ratios (OR) were as follows: poorest (OR: 0.29, 95% CI: 0.24−0.36), poor (OR: 0.43, 95% CI: 0.35−0.53), middle (OR: 0.34, 95% CI: 0.26−0.43), richer (OR: 0.42, 95% CI: 0.33−0.53), and richest (OR: 0.52, 95% CI: 0.40−0.67).

**Table 5.** Multivariate logistic regression on full immunization among children 12-23 months by household wealth status, India, 2019-21

| **Background characteristic** | **Poorest**<br>**OR, [95%,**<br>**C.I]** | **Poor**<br>**OR, [95%,**<br>**C.I]** | **Middle**<br>**OR, [95%,**<br>**C.I]** | **Richer**<br>**OR, [95%,**<br>**C.I]** | **Richest**<br>**OR, [95%,**<br>**C.I]** |
| --- | --- | --- | --- | --- | --- |
| **Recent Migration (<3 years)** |  |  |  |  |  |
| No<sup>Ref</sup> | 1 | 1 | 1 | 1 | 1 |
| Yes | 0.29*** | 0.43*** | 0.34*** | 0.42*** | 0.52*** |
|  | [0.24,0.36] | [0.35,0.53] | [0.26,0.43] | [0.33,0.53] | [0.40,0.67] |
| **Gender** |  |  |  |  |  |
| Male<sup>Ref</sup> | 1 | 1 | 1 | 1 | 1 |
| Female | 0.90<br>[0.81,1.01] | 0.94<br>[0.83,1.06] | 1.05<br>[0.91,1.21] | 0.93<br>[0.78,1.11] | 1.04<br>[0.87,1.25] |
| **Mother's age at birth** |  |  |  |  |  |
| 15-19 years<sup>Ref</sup> | 1 | 1 | 1 | 1 | 1 |
| 20-29 years | 1.26*<br>[1.04,1.52] | 1.26*<br>[1.01,1.56] | 1.00<br>[0.78,1.28] | 1.28<br>[0.94,1.72] | 0.78<br>[0.48,1.24] |
| 30+years | 1.39**<br>[1.08,1.78] | 2.06***<br>[1.51,2.81] | 1.28<br>[0.92,1.78] | 0.97<br>[0.64,1.45] | 0.79<br>[0.46,1.36] |
| **Mother's education in years** |  |  |  |  |  |
| No schooling<sup>Ref</sup> | 1 | 1 | 1 | 1 | 1 |
| <5 years complete | 1.52***<br>[1.22,1.90] | 1.50**<br>[1.12,2.00] | 1.47<br>[0.99,2.17] | 1.22<br>[0.69,2.17] | 1.18<br>[0.42,3.33] |
| 5-8 years complete | 1.43***<br>[1.25,1.64] | 1.43***<br>[1.21,1.70] | 1.42**<br>[1.12,1.81] | 1.31<br>[0.95,1.80] | 1.03<br>[0.62,1.73] |
| 9-12 years | 1.58***<br>[1.34,1.87] | 1.46***<br>[1.22,1.75] | 1.59***<br>[1.25,2.02] | 1.41*<br>[1.04,1.93] | 1.05<br>[0.64,1.71] |
| More >12 years | 1.44<br>[0.95,2.21] | 1.72**<br>[1.24,2.37] | 1.54**<br>[1.14,2.08] | 1.16<br>[0.81,1.66] | 0.84<br>[0.51,1.38] |
| **Media Exposure** |  |  |  |  |  |
| No<sup>Ref</sup> | 1 | 1 | 1 | 1 | 1 |
| Yes | 1.31**<br>[1.10,1.55] | 1.17*<br>[1.02,1.34] | 1.24**<br>[1.06,1.46] | 1.04<br>[0.86,1.25] | 1.14<br>[0.91,1.44] |
| **Birth Order** |  |  |  |  |  |
| 1<sup>st</sup> order<sup>Ref</sup> | 1 | 1 | 1 | 1 | 1 |
| 2 & 3<sup>rd</sup> order | 0.78**<br>[0.67,0.91] | 0.81**<br>[0.70,0.94] | 0.80**<br>[0.67,0.95] | 0.90<br>[0.74,1.09] | 0.77**<br>[0.63,0.94] |
| 4<sup>th</sup> & more orders | 0.69***<br>[0.57,0.84] | 0.64**<br>[0.49,0.84] | 0.58***<br>[0.44,0.76] | 0.73<br>[0.51,1.04] | 0.60*<br>[0.38,0.96] |
| **Intended Birth** |  |  |  |  |  |
| Yes<sup>Ref</sup> | 1 | 1 | 1 | 1 | 1 |
| No | 0.95<br>[0.79,1.15] | 0.97<br>[0.77,1.22] | 0.81<br>[0.55,1.18] | 0.93<br>[0.68,1.27] | 0.80<br>[0.58,1.11] |
| **Institutional delivery** |  |  |  |  |  |
| Public Health Facility<sup>Ref</sup> | 1 | 1 | 1 | 1 | 1 |
| Private Health Facility | 0.67***<br>[0.54,0.81] | 0.85<br>[0.72,1.01] | 0.80*<br>[0.67,0.95] | 0.85<br>[0.70,1.03] | 0.85<br>[0.70,1.04] |
| No Health Facility | 0.63***<br>[0.55,0.72] | 0.59***<br>[0.49,0.72] | 0.62***<br>[0.48,0.80] | 0.81<br>[0.53,1.24] | 0.51*<br>[0.30,0.86] |
| **Current place of residence** |  |  |  |  |  |
| Urban<sup>Ref</sup> | 1 | 1 | 1 | 1 | 1 |
| Rural | 1.54**<br>[1.14,2.10] | 1.42**<br>[1.13,1.78] | 1.30**<br>[1.07,1.56] | 1.04<br>[0.87,1.26] | 1.21*<br>[1.02,1.44] |
| **Social Groups** |  |  |  |  |  |
| Schedule caste<sup>Ref</sup> | 1 | 1 | 1 | 1 | 1 |
| Schedule tribe | 1.13<br>[0.96,1.33] | 1.27*<br>[1.01,1.60] | 1.01<br>[0.74,1.39] | 0.87<br>[0.40,1.86] | 0.67<br>[0.38,1.18] |
| OBC | 0.93<br>[0.80,1.07] | 1.01<br>[0.86,1.18] | 0.93<br>[0.77,1.13] | 1.22<br>[0.97,1.54] | 1.22<br>[0.94,1.59] |
| Others | 0.98<br>[0.78,1.21] | 1.08<br>[0.85,1.37] | 0.88<br>[0.69,1.12] | 0.99<br>[0.76,1.28] | 0.93<br>[0.71,1.21] |
| **Religion** |  |  |  |  |  |
| Hindu<sup>Ref</sup> | 1 | 1 | 1 | 1 | 1 |
| Muslim | 0.72***<br>[0.59,0.87] | 0.94<br>[0.76,1.14] | 0.82<br>[0.67,1.01] | 0.67***<br>[0.53,0.84] | 0.81<br>[0.62,1.06] |
| Christian | 0.81<br>[0.60,1.09] | 0.77<br>[0.51,1.18] | 1.37<br>[0.81,2.35] | 1.42<br>[0.77,2.62] | 2.25**<br>[1.23,4.12] |
| Sikh | 0.33<br>[0.07,1.52] | 0.92<br>[0.46,1.86] | 0.67<br>[0.40,1.15] | 0.83<br>[0.55,1.25] | 0.92<br>[0.65,1.29] |
| Others | 1.56*<br>[1.01,2.42] | 0.94<br>[0.46,1.93] | 1.97<br>[0.83,4.70] | 1.80<br>[0.66,4.91] | 0.94<br>[0.34,2.60] |
| **Regions of India** |  |  |  |  |  |
| North<sup>Ref</sup> | 1 | 1 | 1 | 1 | 1 |
| Central | 0.71**<br>[0.57,0.89] | 0.74**<br>[0.60,0.91] | 0.81*<br>[0.66,0.99] | 0.72**<br>[0.58,0.91] | 0.64***<br>[0.52,0.79] |
| East | 1.14<br>[0.91,1.43] | 1.41**<br>[1.12,1.78] | 1.36*<br>[1.06,1.73] | 0.92<br>[0.68,1.24] | 0.74<br>[0.55,1.01] |
| Northeast | 0.57***<br>[0.44,0.74] | 0.49***<br>[0.38,0.64] | 0.54***<br>[0.39,0.75] | 0.52**<br>[0.34,0.81] | 0.53*<br>[0.30,0.94] |
| West | 0.85<br>[0.61,1.19] | 0.64**<br>[0.47,0.87] | 0.75<br>[0.56,1.01] | 0.69*<br>[0.51,0.94] | 0.89<br>[0.64,1.23] |
| South | 0.89<br>[0.57,1.41] | 1.23<br>[0.92,1.63] | 1.20<br>[0.93,1.56] | 1.18<br>[0.92,1.51] | 0.84<br>[0.64,1.10] |
| **Sample** | **11289** | **9988** | **8468** | **7400** | **5987** |
| Note- * p<0.05, ** p<0.01, *** p<0.001 |  |  |  |  |  |

Table 6 presents the relationship between complete immunization and children’s migration status aged 12-23 months, categorized by social group. The results indicate lower odds of full immunization among children in all social groups who have recently migrated. The odds ratios were as follows: Schedule caste (OR: 0.35, 95% CI: 0.28−0.42), Schedule tribe (OR: 0.48, 95% CI: 0.34−0.69), Other Backward Classes (OR: 0.40, 95% CI: 0.35−0.46), and Others (OR: 0.38, 95% CI: 0.30−0.48) compared with non-migrated children in all social groups.

**Table 6.** Multivariate logistic regression on Full immunization among children 12-23 months by mother social groups, India, 2019-21

| **Background characteristic** | **Schedule caste** | **Schedule tribe** | **OBC** | **Others** |
| --- | --- | --- | --- | --- |
| **Recent Migration (<3 years)** | **OR, [95%, C.I]** | **OR, [95%, C.I]** | **OR, [95%, C.I]** | **OR, [95%, C.I]** |
| No<sup>Ref</sup> | 1 | 1 | 1 | 1 |
| Yes | 0.35***<br>[0.28,0.42] | 0.48***<br>[0.34,0.69] | 0.40***<br>[0.35,0.46] | 0.38***<br>[0.30,0.48] |
| **Gender** |  |  |  |  |
| Male<sup>Ref</sup> | 1 | 1 | 1 | 1 |
| Female | 0.86* [0.76,0.99] | 1.03 [0.86,1.24] | 0.98 [0.89,1.07] | 1.01 [0.88,1.17] |
| **Mother's age at birth** |  |  |  |  |
| 15-19 years<sup>Ref</sup> | 1 | 1 | 1 | 1 |
| 20-29 years | 1.38**<br>[1.11,1.72] | 1.13 [0.82,1.56] | 1.08 [0.92,1.28] | 1.12 [0.88,1.44] |
| 30+years | 2.02***<br>[1.51,2.70] | 1.08 [0.67,1.72] | 1.10 [0.88,1.37] | 1.18 [0.86,1.62] |
| **Mother's education in years** |  |  |  |  |
| No schooling<sup>Ref</sup> | 1 | 1 | 1 | 1 |
| <5 years complete | 1.50* [1.08,2.09] | 1.41* [1.03,1.93] | 1.28* [1.00,1.64] | 1.67**<br>[1.16,2.40] |
| 5-8 years complete | 1.33**<br>[1.12,1.58] | 1.35**<br>[1.08,1.70] | 1.43***<br>[1.24,1.64] | 1.40**<br>[1.12,1.76] |
| 9-12 years | 1.51***<br>[1.24,1.82] | 1.60***<br>[1.25,2.05] | 1.53***<br>[1.32,1.77] | 1.39**<br>[1.09,1.76] |
| More >12 years | 1.22 [0.92,1.60] | 0.70 [0.38,1.30] | 1.33**<br>[1.11,1.60] | 1.38* [1.03,1.83] |
| **Media Exposure** |  |  |  |  |
| No<sup>Ref</sup> | 1 | 1 | 1 | 1 |
| Yes | 1.25**<br>[1.08,1.46] | 1.18 [0.96,1.46] | 1.22***<br>[1.09,1.37] | 1.04 [0.89,1.22] |
| **Birth Order** |  |  |  |  |
| 1<sup>st</sup> order<sup>Ref</sup> | 1 | 1 | 1 | 1 |
| 2 & 3<sup>rd</sup> order | 0.74***<br>[0.63,0.87] | 0.72**<br>[0.58,0.90] | 0.89* [0.79,0.99] | 0.80* [0.68,0.95] |
| 4<sup>th</sup> & more orders | 0.61***<br>[0.48,0.78] | 0.73* [0.55,0.97] | 0.72***<br>[0.60,0.86] | 0.74 [0.54,1.01] |
| **Intended Birth** |  |  |  |  |
| Yes<sup>Ref</sup> | 1 | 1 | 1 | 1 |
| No | 0.83 [0.64,1.07] | 0.96 [0.69,1.33] | 0.97 [0.81,1.17] | 0.88 [0.69,1.12] |
| **Institutional delivery** |  |  |  |  |
| Public Health Facility<sup>Ref</sup> | 1 | 1 | 1 | 1 |
| Private Health Facility | 0.81* [0.68,0.97] | 0.60**<br>[0.42,0.86] | 0.81***<br>[0.72,0.91] | 0.83* [0.69,1.00] |
| No Health Facility | 0.78* [0.63,0.95] | 0.46***<br>[0.38,0.56] | 0.63***<br>[0.54,0.74] | 0.60***<br>[0.47,0.76] |
| **Current place of residence** |  |  |  |  |
| Urban<sup>Ref</sup> | 1 | 1 | 1 | 1 |
| Rural | 1.40**<br>[1.14,1.71] | 1.08 [0.69,1.67] | 1.09 [0.95,1.25] | 1.45***<br>[1.20,1.74] |
| **Wealth quintile** |  |  |  |  |
| Poorest<sup>Ref</sup> | 1 | 1 | 1 | 1 |
| Poorer | 1.06 [0.89,1.27] | 1.04 [0.84,1.29] | 1.16* [1.01,1.33] | 1.41**<br>[1.11,1.77] |
| Middle | 1.43**<br>[1.14,1.78] | 1.27 [0.93,1.72] | 1.33***<br>[1.14,1.55] | 1.58***<br>[1.23,2.03] |
| Richer | 1.33* [1.03,1.72] | 1.06 [0.62,1.81] | 1.46***<br>[1.22,1.75] | 1.59**<br>[1.19,2.13] |
| Richest | 1.41* [1.02,1.95] | 1.11 [0.59,2.10] | 1.42**<br>[1.15,1.76] | 1.68**<br>[1.21,2.34] |
| **Religion** |  |  |  |  |
| Hindu<sup>Ref</sup> | 1 | 1 | 1 | 1 |
| Muslim | 0.58**<br>[0.40,0.82] | 0.91 [0.50,1.64] | 0.75***<br>[0.66,0.85] | 0.90 [0.76,1.07] |
| Christian | 1.00 [0.59,1.69] | 1.02 [0.72,1.43] | 2.34* [1.12,4.88] | 2.19* [1.10,4.36] |
| Sikh | 0.57**<br>[0.40,0.82] | 0.55 [0.10,2.92] | 2.01* [1.09,3.72] | 1.06 [0.73,1.55] |
| Others | 1.07 [0.55,2.11] | 1.88**<br>[1.24,2.84] | 0.42 [0.12,1.46] | 3.64**<br>[1.64,8.06] |
| **Regions of India** |  |  |  |  |
| North<sup>Ref</sup> | 1 | 1 | 1 | 1 |
| Central | 0.75**<br>[0.61,0.92] | 0.67**<br>[0.50,0.89] | 0.78**<br>[0.67,0.91] | 0.63***<br>[0.51,0.76] |
| East | 1.11 [0.88,1.39] | 1.17 [0.86,1.60] | 1.16 [0.97,1.38] | 1.28* [1.02,1.61] |
| Northeast | 0.45***<br>[0.31,0.67] | 0.43***<br>[0.30,0.62] | 0.67**<br>[0.51,0.88] | 0.53***<br>[0.41,0.67] |
| West | 0.59**<br>[0.41,0.84] | 0.83 [0.56,1.23] | 0.87 [0.70,1.08] | 0.69**<br>[0.53,0.91] |
| South | 1.31* [1.00,1.72] | 0.73 [0.48,1.11] | 1.17 [0.97,1.41] | 0.80 [0.60,1.06] |
| **Sample** | **8881** | **8495** | **16603** | **9153** |
| Note- * p<0.05, ** p<0.01, *** p<0.001 |  |  |  |  |

## Discussion

This study investigated the relationship between maternal/parental migration and children’s full immunization coverage by their first birthday. Recent migrants were defined as children with mothers residing in the current location for less than 3 years, including visitors who temporarily stayed at the surveyed households prior to the survey. The study revealed five significant findings regarding the full immunization of children and migration in India. First, 9.6% of children had recently migrated to their current residence (within the past 3 years). Second, based on recent migration status, disparities in full immunization coverage were observed across major states, among socio-economic groups, and for children of mothers with lower education levels. Third, recently migrated children were less likely to receive complete immunization than non-migrated children, even after excluding data from children whose mothers were not living with their husbands, husbands with multiple wives, and visitors. Additionally, children who were recently migrated were less likely to be fully immunized than both permanent residents and settled migrants. Fourth, a consistent finding was that recently migrated children, across household wealth quintiles (poorest, poor, middle, richer, and richest), had a lower probability of achieving full immunization than their non-migrated counterparts. Fifth, similar disparities were observed across different social groups, with recently migrated children in these groups also showing a lower likelihood of full immunization than non-migrated children.

Numerous studies have demonstrated that demographic, socio-economic, and geographic factors significantly influence vaccination rates. However, there is limited evidence on the relationship between migration and childhood vaccination in low-income countries, including India. Our study indicated that recent migration of less than 3 years significantly contributes to lower full immunization uptake among children aged 12-23 months in India. The analysis also highlights that recent migration across wealth status and social groups was associated with low immunization uptake among children aged 12-23 months. The existing studies support our findings, showing that children with recent migrant status are less likely to be fully immunized than natives and settled migrants [19,35]. These studies also define recent migration, similar to our study, as those who have lived at their current residence for at least 2 years. Kusuma et al. (2010, 2018) found that recent migrants (those who moved to the national capital, Delhi, from rural areas within the last 5 years) had a higher risk of incomplete immunization than settled migrants [18,28]. However, studies that did not consider recent migration, such as living in the current place for less than 3 years, were also found to be associated with lower routine immunization coverage among children. Kiros & White (2004) and Antai D. (2010) found that children who migrated from rural to urban areas were less likely to be fully immunized than children of non-migrant mothers residing in rural areas, even after adjusting for several child, family, and village-level variables [23,36].. In addition, evidence from systematic reviews and meta-analyses also suggests that migrant children, especially recent migrants, are less likely to receive full immunization than non-migrant children [26,37]. Our results demonstrate how recent migration may influence children’s immunization uptake through the health access livelihood framework and the theories of migrant disruption, migrant adaptation, and assimilation. Relocating to a current residential place, even within the same country or state, alters both social and physical environments, profoundly impacting health behaviors and outcomes. For recent migrants, whether moving from rural to urban areas or vice versa, their access to healthcare services is often restricted. This difficulty stems from a combination of factors in the host community. These factors include their living and working conditions, limited employment opportunities, inadequate social security benefits, and their social groups. Collectively, these issues compromise their ability to afford essential healthcare services [14]. Additional specific barriers, including linguistic challenges, prevailing social norms, or cultural perceptions of health, may further restrict migrants’ access to services. Consistent with both our study and the theory of migrant adaptation, Azad and Islam’s study indicates that recent migrants have lower immunization rates than settled migrants [38]. This difference is likely due to an increased familiarity with local cultural practices, norms, and living conditions among settled migrants. In contrast, recent migrants remain more vulnerable, often lacking social networks, which may hinder their access. Poor adaptation also partly explains why migrant children have lower immunization uptake than urban non-migrant children. According to a regional study conducted in India [19], migrant parents’ use of immunization services is negatively affected by their limited knowledge of vaccination schedules and timing, as well as limited access to healthcare. It is also evident that migrant children’s vaccination cards are often misplaced during migration, discouraging parents from immunizing their children and leading to interruptions in vaccination schedules. In addition to the disruption theory, our study also assumes that women often return to their native places (particularly in India and other South Asian countries) during childbirth and the immediate postnatal period for better care [39,40]. Studies from Indian states showed that one to two-thirds of women returned to their natal homes at some point during pregnancy, for childbirth, or postpartum, which can also lead to delays or under immunization in the timely uptake of immunizations for their children.

Migratory individuals and families are considered high-risk groups due to their inherent vulnerabilities, which often result in reduced immunization rates and lower uptake of essential health services. Structural barriers in healthcare systems disproportionately impede access to, affordability, and awareness of essential health services, including immunization. These include challenges such as migrants’ long and inconvenient working hours coinciding with immunization session timings and the risk of losing daily wages, often referred to as the opportunity cost. In addition, migrants often face a lack of culturally and linguistically appropriate health information as well as health providers. Together, this translates into an inability to prioritize healthcare over survival, often leading to delays in service uptake and worsening health conditions. Therefore, it demands continuous efforts to ensure these populations are effectively reached and monitored. With rapidly growing urbanization in India, the projected proportions of urban and rural populations are expected to change rapidly over the next two decades. While the current urbanization rate is approximately 35%, it is projected to reach 46.5% by 2050 [41]. The primary factor contributing to this growing urban population is migration, which consequently strains health systems’ infrastructure and service delivery.

To effectively address these challenges, there is a clear need for a robust, inclusive tracking platform built around a unique, portable identifier, similar to a mobile number, that can track the child regardless of geographic movement. Such a system would enable both caregivers and frontline health workers to access and update vaccination records in real time, ensuring continuity of care even when families migrate. Evidence from multiple studies suggests that the use of unique digital identifiers for tracking migratory children significantly improves immunization coverage by reducing dropouts and missed doses.

In India, nearly 20% of children remain partially immunized, representing a critical “low-hanging fruit” for programmatic intervention, and a substantial proportion of these children belong to migrant populations. Additionally, zero-dose children, who account for roughly 4% of the cohort, also disproportionately come from migratory backgrounds. Frequent movement during early childhood disrupts immunization schedules, often compounded by limited awareness and weak linkages to local health systems. Addressing this gap requires a robust beneficiary-tracking mechanism that transcends administrative boundaries. A unified digital system with a unique ID can ensure that children are identified, tracked, and vaccinated regardless of their previous place of residence. This approach would not only facilitate timely follow-up for missed doses but also strengthen accountability within the system, thereby improving coverage and advancing the IA2030 goal.

The introduction of digital tools has further strengthened India’s capacity to track migratory and dropout cases effectively. U-WIN, a digital platform for vaccine registration, record-keeping, and tracking, built on the successful foundation of CoWIN platform, enables real-time registration, tracking, and follow-up of beneficiaries across multiple locations. It is critical in tracing migratory children, ensuring scheduled dose completion, and tracking beneficiaries. It enables appointment booking, real-time data capture, certificate generation, and follow-up reminders to ensure complete vaccination. This strategy has strengthened the continuum of care. In conclusion, these mechanisms have the potential to address challenges in vaccine uptake and ensure that no child is left behind in immunization.

## Limitations

Although this study reveals important relationships between parental migration and FIC, it is limited by data unavailability. The current data is limited and doesn’t include crucial information on the migratory patterns, specifically rural-to-urban and urban-to-rural movements. Additionally, the effect of parental migration (within and across states) on their children’s immunization status is not addressed. It lacks specific information on where migrants reside within urban settings, such as in slums or at their workplaces. Furthermore, NFHS-5 does not provide information about the reasons behind migration or the origins of previous movements in current residences. The survey does not collect social network details for migrant groups, reasons for delays, or non-utilization of healthcare services. These limitations may affect the study’s outcomes.

## Conclusion

The findings of this study provide significant quantitative evidence that recent migration (less than 3 years) of children is a key factor influencing low coverage and is a predictor of full immunization among children aged 12-23 months in India. Additionally, recent migration was consistently associated with a lower likelihood of full vaccination across household wealth levels and social groups. This study suggests that recently migrated children are a vulnerable subgroup at risk of not receiving all basic vaccinations by their first birthday. This study recommends that policymakers and health service administrators, especially in Maternal and Child Health, aim to include recently migrated children. Nevertheless, additional research on migration and childhood vaccination is needed to better understand mechanisms to enhance immunization coverage and uptake of health services among children.

## Authors Contributors

Conceptualisation: Pritu Dhalaria, Ajay Kumar Verma, and Pawan Kumar. Methodology: Ajay Kumar Verma and Ajeet Kumar Singh. Formal analysis: Ajay Kumar Verma and Ajeet Kumar Singh. Investigation: Pritu Dhalaria, Pretty Priyadarshini, and Kapil Singh. Writing—original draft preparation: Ajay Kumar Verma, Pretty Priyadarshini, and Ajeet Kumar Singh. Writing—review and editing: Ajay Kumar Verma, Pretty Priyadarshini, Ajeet Kumar Singh, Pawan Kumar, and Bhupendra Tripathi. Visualisation: Ajay Kumar Verma, Pretty Priyadarshini and Ajeet Kumar Singh. Supervision: Pritu Dhalaria, Pawan Kumar, Sanjay Kapur, Arindam Ray, and Bhupendra Tripathi. Guarantor information: Ajay Kumar Verma accepts full responsibility for the finished work and/or the conduct of the study, has access to the data, and controls the decision to publish.

## Data availability statement

Data are available in a public, open-access repository. The data are available in the public domain and can be downloaded on reasonable request. https://dhsprogram.com/data/available-datasets.cfm

## Ethics Statement

The study is based on secondary household survey data available in the public domain, with no primary data collection and no human participants involved directly in it; hence, no ethical approval is required.

## Reference

1. Immunization agenda 2030: A global strategy to leave no one behind. Vaccine 42, S5–S14 (2024).

2. Nandi, A. & Shet, A. Why vaccines matter: understanding the broader health, economic, and child development benefits of routine vaccination. Human vaccines & immunotherapeutics 16, 1900–1904 (2020).

3. Zhang, H., Patenaude, B., Zhang, H., Jit, M. & Fang, H. Global vaccine coverage and childhood survival estimates: 1990--2019. Bulletin of the World Health Organization 102, 276 (2024).

4. Shattock, A. J., et al. Contribution of Vaccination to Improved Survival and Health: Modelling 50 Years of the Expanded Programme on Immunization.? The Lancet,?403(10441), 2307-2316. (2024).

5. Li, X., et al. Estimating the Health Impact of Vaccination against Ten Pathogens in 98 Low-Income and Middle-Income Countries from 2000 to 2030: A Modelling Study.? The Lancet,?397(10272), 398-408. (2021).

6. Ministry of Health and Family Welfare. India’s percentage of Zero-dose children to the total population has declined from 0.11% in 2023 to 0.06% in 2024, positioning it as a global exemplar in child health, as acknowledged by the UN Inter-agency Group for Child Mortality Estimation in its 2024 report. Press Information Bureau https://www.pib.gov.in/PressReleasePage.aspx?PRID=2140343&reg=3&lang=2 (2025).

7. Dhalaria, P. et al. Path to full immunisation coverage, role of each vaccine and their importance in the immunisation programme: A cross-sectional analytical study of India. BMJ Public Health 3, (2025).

8. ICF, I. I. for P. S. (IIPS) and. National Family Health Survey (NFHS-5), 2019–21: India. National Family Health Survey, India (2021).

9. Wahl, B. et al. Change in full immunization inequalities in Indian children 12–23 months: an analysis of household survey data. BMC public health 21, 841 (2021).

10. UNICEF. Immunization among Tribal Population in India: A Need Assessment Report-UNICEF. Accessed on Jan 7, 2022. Available from: https://Immunization+among/Tribal. Population/in/India (2022).

11. Kalia, M., Sharma, M., Rohilla, R. & Rana, K. Trend of immunization & gap in vaccine doses as observed in National Family Health Survey rounds in India. The Indian Journal of Medical Research 160, 303 (2024).

12. Panda, B. K., Kumar, G. & Mishra, S. Understanding the full-immunization gap in districts of India: a geospatial approach. Clinical Epidemiology and Global Health 8, 536–543 (2020).

13. Singh, S. K. & Vishwakarma, D. Spatial heterogeneity in the coverage of full immunization among children in India: Exploring the contribution of immunization card. Children and Youth Services Review 121, 105701 (2021).

14. Assaf, S., Thapa, N. R. & Edmeades, J. Internal Adult Women Migrants’ Use and Access to Health Services in 15 DHS Countries. (2023).

15. Cherri, Z. et al. The immune status of migrant populations in Europe and implications for vaccine-preventable disease control: a systematic review and meta-analysis. Journal of Travel Medicine 31, taae033 (2024).

16. Deal, A. et al. Migration and outbreaks of vaccine-preventable disease in Europe: a systematic review. The Lancet Infectious Diseases 21, e387–e398 (2021).

17. Bouaddi, O., et al. Vaccination coverage and access among children and adult migrants and refugees in the Middle East and North African region: a systematic review and meta-analysis. EClinicalMedicine 78, (2024).

18. Kusuma, Y. S., Kaushal, S., Sundari, A. B. & Babu, B. V. Access to childhood immunisation services and its determinants among recent and settled migrants in Delhi, India. Public Health 158, 135–143 (2018).

19. Savani, N. M., Gurjar, Y., Lodha, N. & Nathwani, R. Uptake of Immunization and Its Determinants among Children of Migrant Population Residing in Amreli, Gujarat, India. Pediatric Infectious Disease 5, 114–119 (2023).

20. Government of India. Annual Report, Periodic Labour Force Survey (PLFS), July 2021 - June 2022. https://www.mospi.gov.in/sites/default/files/publication_reports/AnnualReportPLFS2021-22F1.pdf (2022).

21. Ellaway, A., Macintyre, S. & MacKay, L. Migration and Health: A Review of the International Literature. (MRC Social and Public Health Sciences Unit, 2003).

22. Wickramage, K., Vearey, J., Zwi, A. B., Robinson, C. & Knipper, M. Migration and health: a global public health research priority. BMC public health 18, 987 (2018).

23. Antai, D. Migration and child immunization in Nigeria: individual-and community-level contexts. BMC public health 10, 116 (2010).

24. Lindstrom, D. P. & Saucedo, S. G. The short-and long-term effects of US migration experience on Mexican women’s fertility. Social Forces 80, 1341–1368 (2002).

25. Andersen, R. M. Revisiting the behavioral model and access to medical care: does it matter? Journal of health and social behavior 1–10 (1995).

26. Awoh, A. B. & Plugge, E. Immunisation coverage in rural–urban migrant children in low and middle-income countries (LMICs): a systematic review and meta-analysis. J Epidemiol Community Health 70, 305–311 (2016).

27. Fiedler, J. L. A review of the literature on access and utilization of medical care with special emphasis on rural primary care. Social Science & Medicine. Part C: Medical Economics 15, 129–142 (1981).

28. Kusuma, Y. S., Kumari, R., Pandav, C. S. & Gupta, S. K. Migration and immunization: determinants of childhood immunization uptake among socioeconomically disadvantaged migrants in Delhi, India. Tropical Medicine & International Health 15, 1326–1332 (2010).

29. Thomson, A., Robinson, K. & Vallée-Tourangeau, G. The 5As: A practical taxonomy for the determinants of vaccine uptake. Vaccine 34, 1018–1024 (2016).

30. Crawshaw, A. F. et al. Defining the determinants of vaccine uptake and undervaccination in migrant populations in Europe to improve routine and COVID-19 vaccine uptake: a systematic review. The Lancet Infectious Diseases 22, e254–e266 (2022).

31. Smedley, J. et al. Influenza immunisation: attitudes and beliefs of UK healthcare workers. Occupational and environmental medicine 64, 223–227 (2007).

32. Prusty, R. K. & Keshri, K. Differentials in child nutrition and immunization among migrants and non-migrants in urban India. International Journal of Migration, Health and Social Care 11, 194–205 (2015).

33. Mishra, P. S., Choudhary, P. K. & Anand, A. Migration and child health: Understanding the coverage of child immunization among migrants across different socio-economic groups in India. Children and youth services review 119, 105684 (2020).

34. Geddam, J. B. et al. Immunization uptake and its determinants among the internal migrant population living in nonnotified slums of Hyderabad city, India. Journal of family medicine and primary care 7, 796–803 (2018).

35. Hu, Y., Li, Q., Chen, E., Chen, Y. & Qi, X. Determinants of childhood immunization uptake among socio-economically disadvantaged migrants in East China. International journal of environmental research and public health 10, 2845–2856 (2013).

36. Kiros, G.-E. & White, M. J. Migration, community context, and child immunization in Ethiopia. Social Science & Medicine 59, 2603–2616 (2004).

37. Charania, N. A., Gaze, N., Kung, J. Y. & Brooks, S. Vaccine-preventable diseases and immunisation coverage among migrants and non-migrants worldwide: A scoping review of published literature, 2006 to 2016. Vaccine 37, 2661–2669 (2019).

38. Islam, M. M. & Azad, K. M. A. K. Rural--urban migration and child survival in urban Bangladesh: Are the urban migrants and poor disadvantaged? Journal of biosocial science 40, 83–96 (2008).

39. Diamond-Smith, N. G. et al. Defining and characterizing temporary childbirth migration in India. Population research and policy review 44, 1–19 (2025).

40. El Ayadi, A. M., et al. Temporary Childbirth Migration in India: Decision-Making, Decision-Makers, and Influencing Factors. (2025).

41. S Irudaya Rajan & J Retnakumar. UNRAVELLING INDIA’S DEMOGRAPHIC FUTURE Population Projections for States and Union Territories 2021–2051. file:///C:/Users/averma1/Downloads/PFI-and-IIMAD.pdf (2025).

